# Genetic regulation of right ventricular trabeculation identifies developmental and cardiopulmonary pathways

**DOI:** 10.64898/2026.07.24.26358843

**Authors:** Kathryn A McGurk, Jan Sedlacik, Riyad Janan, Arunashis Sau, Fu Siong Ng, Wenjia Bai, James S Ware, Declan P O’Regan

**Affiliations:** National Heart and Lung Institute, Imperial College London, London, UK; MRC Laboratory of Medical Sciences and Institute of Clinical Sciences, Imperial College London, London, UK; Program in Medical and Population Genetics, The Broad Institute of MIT and Harvard, Cambridge, US; Department of Cardiology, Imperial College Healthcare NHS Trust, London, UK; Chelsea and Westminster Hospital NHS Foundation Trust, London, UK; Department of Computing, Imperial College London, London, UK; Department of Brain Sciences, Imperial College London, London, UK; Royal Brompton & Harefield Hospitals, Guy’s and St. Thomas’ NHS Foundation Trust, London, UK

## Abstract

Right ventricular (RV) trabeculation may reflect adaptation to loading conditions and imprinting of early developmental processes, yet its genetic determinants and clinical relevance in adults remain poorly understood. In contrast to the left ventricle, the RV has distinct developmental origins, geometry and loading conditions, suggesting chamber-specific mechanisms of trabecular remodelling. Using deep learning–based image segmentation and fractal dimension analysis, we quantified RV trabecular complexity in diastole and systole in 48,118 UK Biobank participants with genetic data. RV trabecular morphology was associated with systolic function, as well as cardiometabolic and respiratory conditions. Genetic analyses across the allele frequency spectrum identified 52 common loci and 45 genes with a burden of protein-altering variants, implicating sarcomeric function, cytoskeletal organisation, and early cardiac patterning. Although many loci showed shared effects across both ventricles, we also identified RV-specific genetic associations linked to congenital heart disease and respiratory phenotypes. Notably, associations at *CFTR* suggest a connection between airway-associated mucus regulation and RV remodelling, whereas *MYH6* implicates sarcomeric and developmental mechanisms in adult RV patterning. Together, these findings establish RV trabeculation as a trait that captures both shared and chamber-specific mechanisms relevant to cardiovascular health and disease.

## Introduction

The ventricular endocardium is lined by a complex network of muscular trabeculae that forms an interface between the compact myocardium and intracardiac blood flow. Trabeculae are essential in early cardiac development, where they facilitate oxygen diffusion before coronary maturation and contribute to conduction system formation, chamber morphogenesis, and ventricular patterning^1–3^. In the adult heart, their function is less well resolved, but growing evidence suggests that trabecular architecture remains biologically relevant to ventricular mechanics, flow dynamics, and myocardial remodelling^4–6^.

Increased trabeculation may arise as a physiological and reversible response to altered loading conditions, such as pregnancy or athletic training^7,8^, and is often benign in healthy individuals without symptoms or evidence of inherited heart disease^9^. Conversely, altered trabecular morphology is also observed in cardiomyopathy, heart failure, and other systemic disorders, where it may carry prognostic information beyond conventional imaging and electrocardiographic markers^9–16^. Together, these findings support a model in which trabeculation reflects both developmental patterning and adaptive remodelling in response to haemodynamic and myocardial stress.

We previously showed that LV trabecular morphology is associated with cardiomyopathies, valve and conduction disease, and is shaped by both common and rare genetic variation^17^. We also found that changes in LV trabeculation may occur early in the natural history of heart disease, supporting its use as a marker of remodelling and a route to identifying disease modifiers^17,18^. Whether these principles extend to the right ventricle remains unknown.

This question is important because the RV is developmentally, functionally, and morphologically distinct from the LV. RV hypertrabeculation has been described in association with congenital heart disease, heart failure, and arrhythmias^19^. However, the extent to which RV trabecular morphology reflects chamber-specific biology, relates to cardiopulmonary and metabolic disease in the general population, or is governed by distinct genetic mechanisms is not known.

Here, we examine the genetic basis of RV trabeculation. We applied deep learning–based image segmentation and fractal dimension analysis to quantify RV trabecular complexity in diastole and systole in 48,118 UK Biobank participants with genetic data. By integrating imaging, phenotypic, and genetic analyses across the allele frequency spectrum, we show that RV trabeculation reflects both shared and right-heart-specific biology with relevance to development, remodelling, and disease (**Figure 1**).

**Figure 1.**
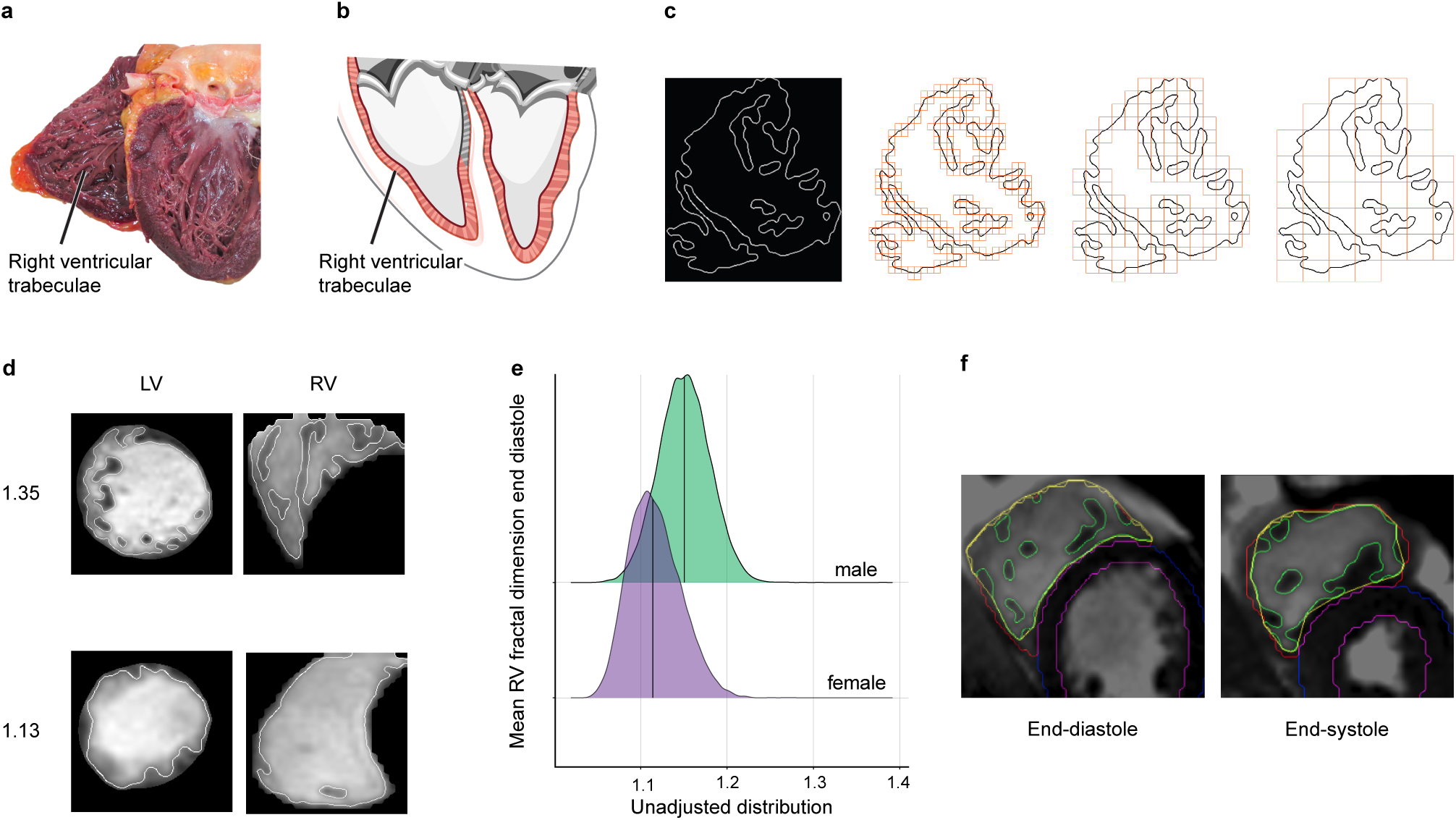
Population right ventricular cardiac trabeculation analysis summary. **a-b**, Myocardial trabeculae on the endocardial surface of the human heart with the right ventricle (RV) noted in the schematics. c, The myocardium was segmented using deep learning algorithms and edge detection was used to define the trabecular boundary in systole and diastole. Trabecular complexity was defined by measuring the fractal dimension (FD) of this boundary using a box-counting methodology. d, CMR was acquired in the short-axis plane, and trabecular morphology was quantified by FD analysis, with examples of FD measured at 1.13 and 1.35 shown for both LV and RV. e, The distribution of raw mean FD values in the RV in end-diastole by sex. f, Example of the resulting FD output with coloured contour lines. FD, fractal dimension; RV, right ventricle; ED, end-diastole; LV, left ventricle; ES, end-systole. The images have been reproduced with permission from the UK Biobank ©. All images show an adult human heart. Image credit: a, Arpatsara/Shutterstock.com; b, GraphicsRF/Shutterstock.com.

## Results

### Study overview

The UK Biobank study recruited 500,000 participants aged 40 to 69 years in the United Kingdom between 2006 and 2010^20^. A sub-study recalled participants for cardiac magnetic resonance imaging (CMR)^21^, and volumetric traits were measured using quality-controlled deep learning algorithms^22–24^. Imaging was made available in two releases. Genotyping array data and exome sequencing data were available for 470,000 participants.

Trabecular morphology was quantified using edge detection of the endocardium to derive a scale-invariant fractal dimension (FD) ratio for each of the RV slices, where a higher value indicates a greater degree of surface complexity. This approach provides a measure of the complexity of the trabecular mesh-work^6,25^. We adapted our previous methods^17^ to look for contrasts between trabecular morphology observed in end-diastole (9 basal to apical image slices) and end-systole (7 slices) and between FD measured in the left (LV) and right ventricles (RV; **Figure 1, Figure S1**)^26^.

The traits were adjusted using multiple linear regression for age at scan, age^2^, sex, age:sex interaction, imaging centre, imaging release, body surface area, systolic blood pressure, vigorous exercise, and 10 genetic principal components of ancestry. Comparisons were also undertaken with and without adjustment for RV volume (end-diastolic, RVEDV; end-systolic, RVESV).

Imaging-derived FD trabecular traits were assessed for association with genetic, phenotypic, and clinical outcome data (**Figure 2**). We analysed the cohort for genome-wide association studies, protein-altering variant burden tests, and curated and phenome-wide association study clinical outcomes. Ancestry, physical activity, electrocardiogram diagnoses, and relationships with CMR-derived traits were also assessed.

**Figure 2.**
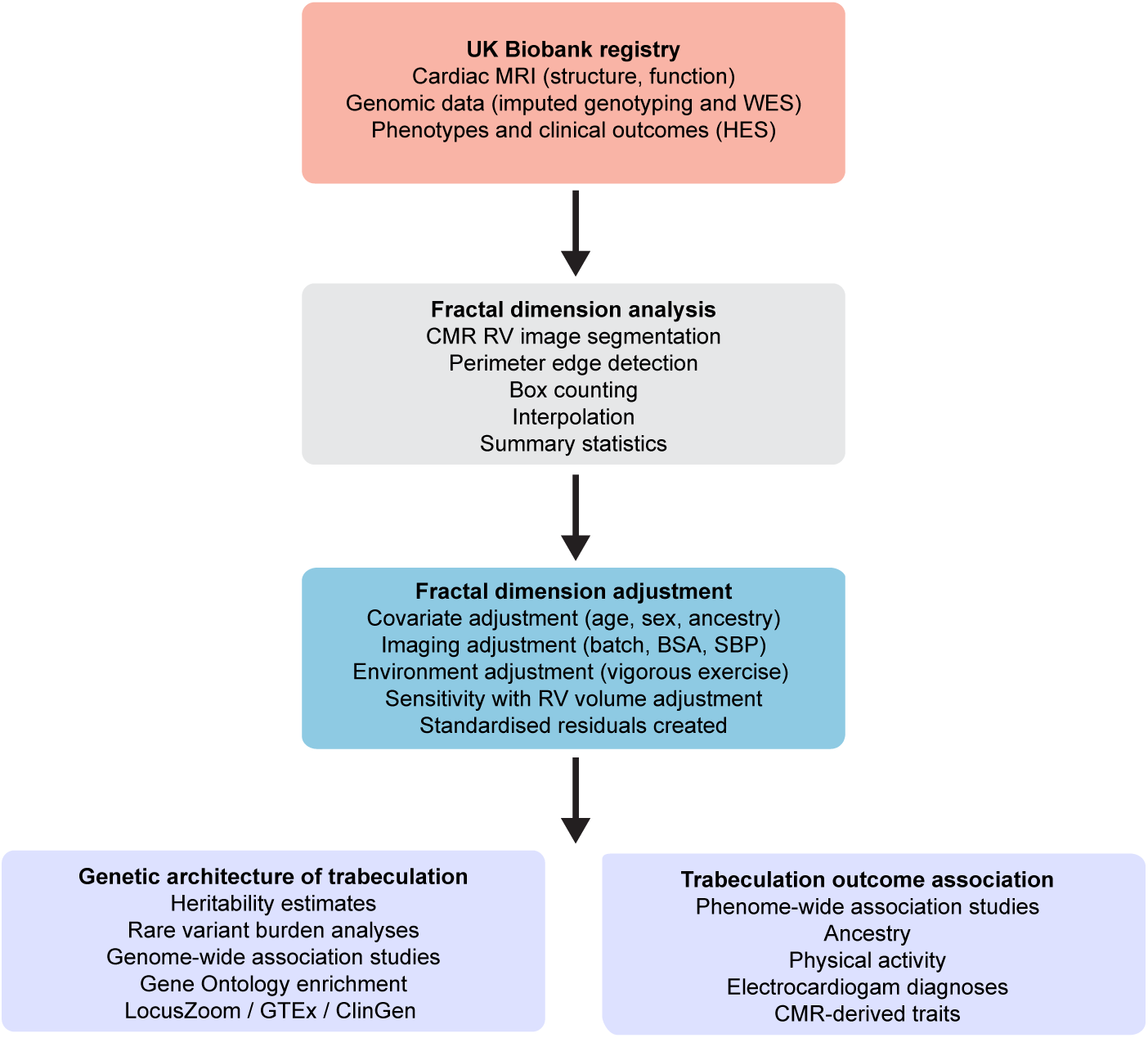
Study flowchart. A summary of the main steps in our analysis of fractal dimension and the genetic and outcome associations. MRI, magnetic resonance imaging; CMR, cardiac MRI; WES, exome sequencing; HES, hospital episode statistics; RV, right ventricle; BSA, body surface area; SBP, systolic blood pressure; GTEx, the Genotype-Tissue Expression (GTEx) Portal; ClinGen, the Clinical Genome Consortium.

### Age, sex, ancestry, imaging-derived traits, and physical activity

Average RV trabeculation is increased with male sex (49%; **Figure S1**), but the relationship with age at MRI was weak (−0.1>R<0.1; average age at scan was 64 years). RV trabeculation was associated with ancestry but with a modest phenotypic effect compared to the LV^17^ and was independent of ventricular volume: African ancestry was associated with increased mean RV FD compared to White British ancestry that dominates the UK Biobank demographics (87%; *β*=0.02). Indian and Chinese ancestry associated with decreased mean RV FD (*β*=-0.01–0.02).

Mean RV FD had the strongest relationships with other CMR-derived measures of RV ventricular volume (enddiastolic [RVEDV R=0.6], end-systolic [RVESV R=0.6], stroke volume [RVSV R=0.4-0.5]; **Figure 3**). Increased mean end-systolic RV FD was associated with decreased RV ejection fraction (RVEF; R=**-**0.5), an inverse relationship that has been described for LV trabecular morphology previously^17,27^.

**Figure 3.**
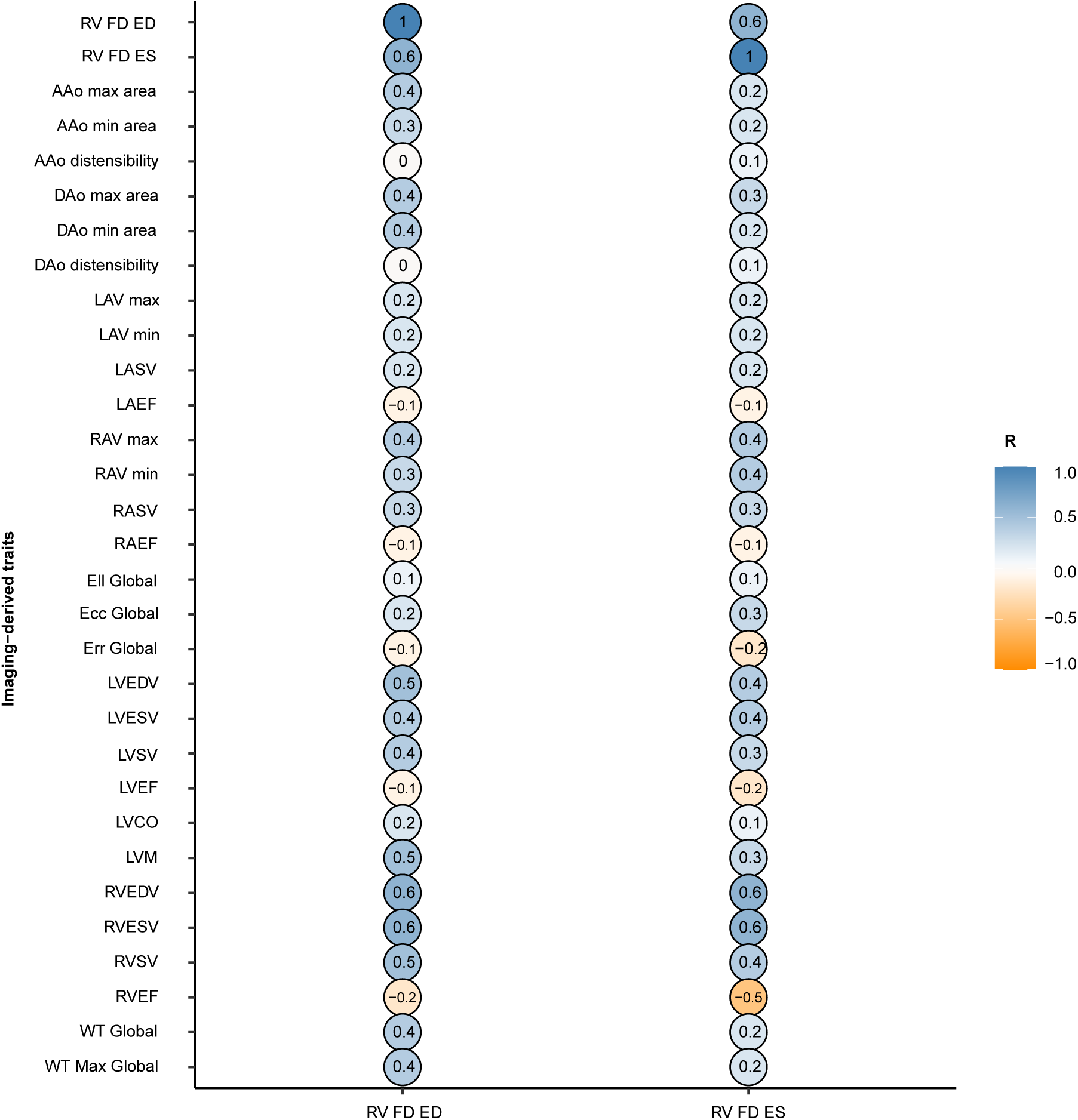
Relationships between imaging-derived traits and right ventricular trabecular morphology. The table quantifies the relationship (correlation coefficient, R) between mean right ventricular fractal dimension in end-diastole and end-systole with two-dimensional summary imaging cardiac measures. RV, right ventricle; FD, fractal dimension; ES, end-systole; ED, end-diastole; EDV, end-diastolic volume; ESV, end-systolic volume; SV, stroke volume; EF, ejection fraction; CO, cardiac output; LAV, left atrial volume; RAV; right atrial volume; AAo, ascending aortic area; DAo, descending aorta area; Ecc, circumferential strain; Err, radial strain; Ell, longitudinal strain; WT, wall thickness.

Physical activity decreased RV FD with increased ventricular volume (beta=-0.22, SE=0.03, P=2.9×10^−12^ comparing 0 days of vigorous activity to 7 days), the relationship between exercise and RV dilatation has been described previously^28^.

### RV trabeculation is altered with circulatory, respiratory, and metabolic diagnoses

Phenome-wide association studies (PheWASs) identified significant associations between measures of RV trabecular morphology and disease-related clinical features (**Figure 4, Figures S2-S5, Table S1**). To assess for pleiotropy with current diagnostic imaging measures, we analysed PheWAS of right heart traits: ventricular end-diastolic volume (RVEDV), end-systolic volume (RVESV), ejection fraction (RVEF), stroke volume (RVSV), and atrial ejection fraction and stroke volume (RAEF, RASV; **Figure 4**).

**Figure 4.**
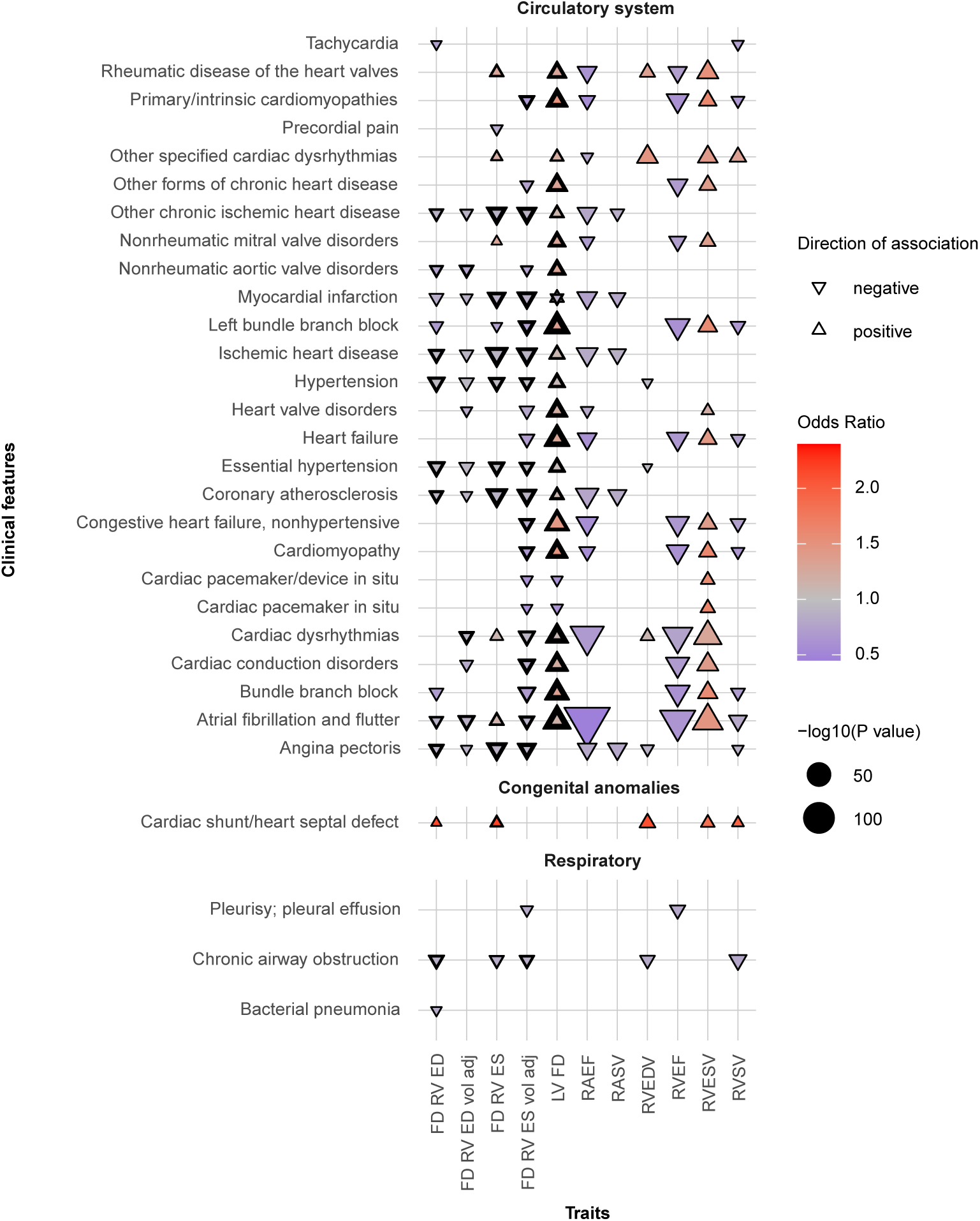
Phenome-wide association study of right ventricular trabecular complexity and other CMR-derived traits. Trabecular morphology was analysed through fractal dimension analysis (FD) of the right ventricle (RV) in end-diastole (ED) and end-systole (ES). ED and ES volume-adjusted RV FD traits are presented separately (vol adj). FD of the left ventricle (LV FD) was added for comparison. Right ventricular end-diastolic volume (RVEDV), end-systolic volume (RVESV), ejection fraction (RVEF), stroke volume (RVSV), and atrial ejection fraction and stroke volume (RAEF, RASV), were analysed to assess for pleiotropy between trabecular complexity (FD) and remodelling traits altered in cardiac disease. Fractal dimension was measured at different spatial locations in the right ventricle, and the aggregate of all results is shown. Only phenotypes that were significant for FD traits are presented. The analyses were completed on 45,681 participants of the UK Biobank population. Phenotypes (phecodes) are shown on the y-axis, with the phecode category separating the groups, and imaging traits are on the x-axis. Each point denotes a significant PheWAS association with a Bonferroni correction for 1,163 analysed phecodes. The shape and colour denote the direction of effect and the odds ratio. Categories and phenotypes other than the circulatory system, congenital anomalies, and respiratory categories, and additional results are presented in the supplementary.

RV FD measures showed associations with disease through increased trabecular complexity for structural heart diseases, but decreased FD for respiratory and metabolic conditions. A positive relationship was found between RV trabecular complexity and RV volume in participants with a cardiac septal defect. RV FD measures had statistically significant inverse relationships (i.e., decreased FD) with respiratory conditions (chronic airway obstruction, bacterial pneumonia, pleural effusion, tobacco use disorder; **Figure 4, Figure S2**). Both the respiratory and septal defect associations were unique to FD measures in the RV and not identified for the LV^17^. Decreased FD was observed for both LV and RV trabecular measures with metabolic conditions: obesity, diabetes, ischemic heart disease, and hyperlipidemia. Cardiomyopathies associated with RV systolic dysfunction and trabeculation: decreased RVEF and RAEF, decreased systolic RV trabeculation, increased RV systolic volume, and increased LV trabeculation, where LV trabeculation has a stronger relationship with cardiomyopathies^17,18^. LV valve diseases showed opposing associations with RV imaging-derived traits: LV mitral valve diseases associated with increased RV volumes, bi-ventricular trabeculation with this increase in volume, and decreased right heart ejection fractions. LV aortic valve diseases associated with decreased RV trabeculation but increased LV trabeculation, independent of volume. A nominal association was observed for more complex FD RV in end systole with rare, non-rheumatic tricuspid valve disorders (ICD 36, n=30 in 42,850, P=0.04, **Figure S6**) but not pulmonary valve disorders (ICD 27-29, 37), or respiratory distress (ICD J80). Of note, measures of FD and thromboembolic events or stroke were not identified at PheWAS.

### Cardiac electrophysiology associates with RV trabeculation and volume

The PheWAS suggested that altered RV trabeculation and volume are associated with underlying electrical remodelling. Associations with RV FD and measures of RV function were identified for UK Biobank baseline electrocardiogram diagnoses^29^ (n=34,346) of first-degree AV block, left bundle branch block, right bundle branch block, sinus bradycardia, sinus tachycardia, and atrial fibrillation (**Figure 5**). The strongest RV signals were observed between sinus bradycardia and a more complex trabecular morphology with increased RV volume. Participants with diagnoses of right bundle branch block had decreased RVEF with increased volumes and trabecular morphology, particularly in systole.

**Figure 5.**
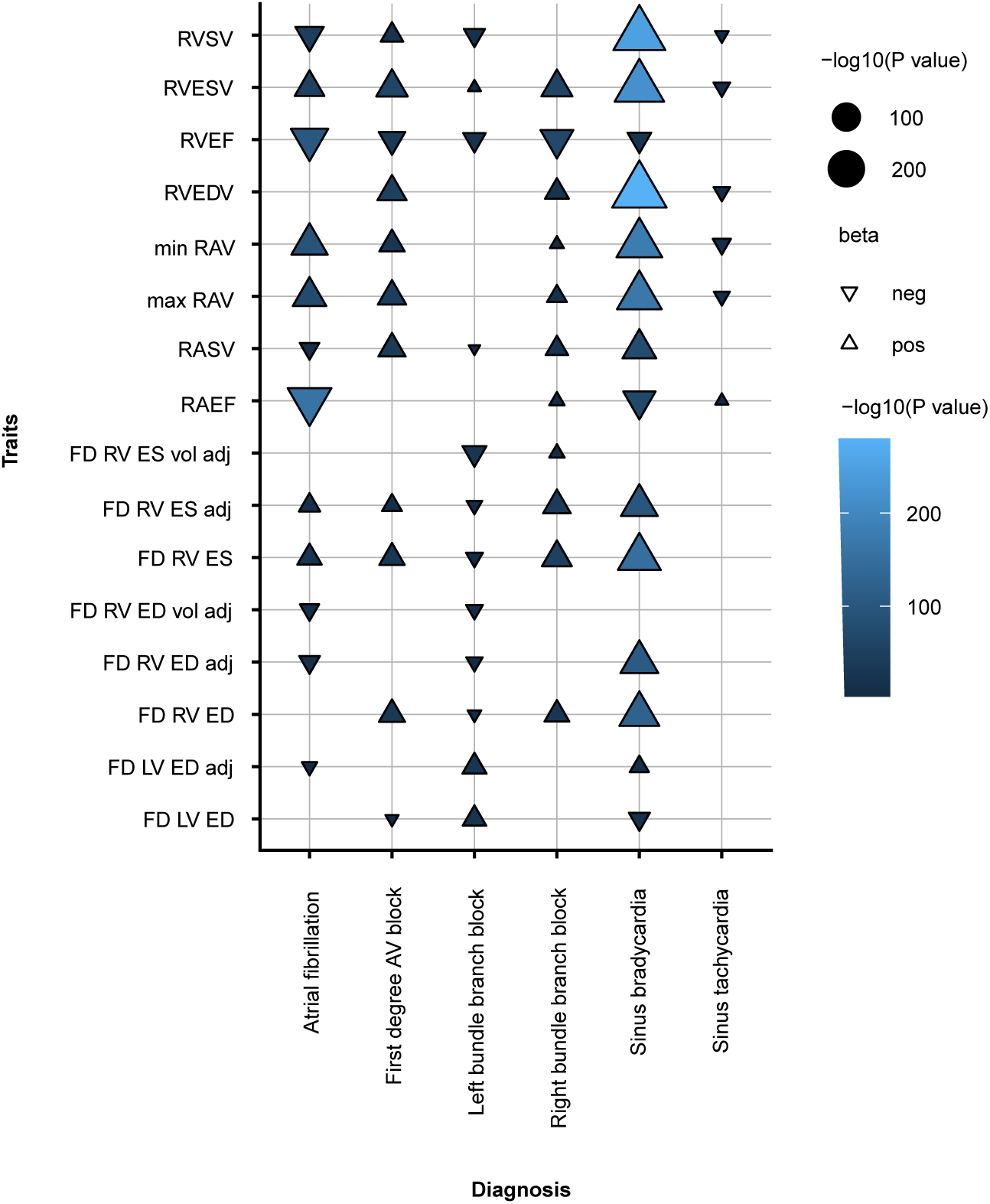
Association study of right heart imaging-derived traits and ECG-derived diagnoses. Trabecular morphology was analysed through fractal dimension analysis (FD) of the right ventricle (RV) in end-diastole (ED) and end-systole (ES). ED and ES covariate-adjusted (adj) and volume-adjusted (vol adj) RV FD traits are presented separately. Right ventricular end-diastolic volume (RVEDV), end-systolic volume (RVESV), ejection fraction (RVEF), stroke volume (RVSV), atrial ejection fraction and stroke volume (RAEF, RASV) were analysed to assess for pleiotropy between trabecular complexity (FD) and remodelling traits altered in cardiac disease. Fractal dimension was measured at different spatial locations in the right ventricle, and the mean measures are shown. The analyses were completed on 34,346 participants of the UK Biobank population. RV traits are described on the y-axis, with the ECG-derived diagnosis on the x-axis. Each point denotes an association; the shape and colour denote the direction of effect and the odds ratio.

### Genome-wide association studies identified genetic loci influencing RV trabeculation

We identified 52 loci from GWASs of 34,110 European participants (**Figure 6, Table S2**). LocusZoom and GTeX eQTLs were used to prioritise genes at each locus. *TBX5, TNNT2*, and *GOSR2* were identified for both ED and ES measures, regardless of volume, and previously for FD measures in the LV (e.g.,^17^), suggesting they are modifiers of bi-ventricular trabecular structure and function. Bi-ventricular genomic loci of *KCNRG, TTN, SFRP1, MECOM, MTSS1, SIPA1L1,* and *ITPR1* were identified from GWAS of the LV also^17^. The loci of genes involved in the regulation of the development of trabeculation and/or cardiomyopathies were identified (e.g., *TBX5, GOSR2, MLF1, TNNT2*)^3,30,31^.

**Figure 6.**
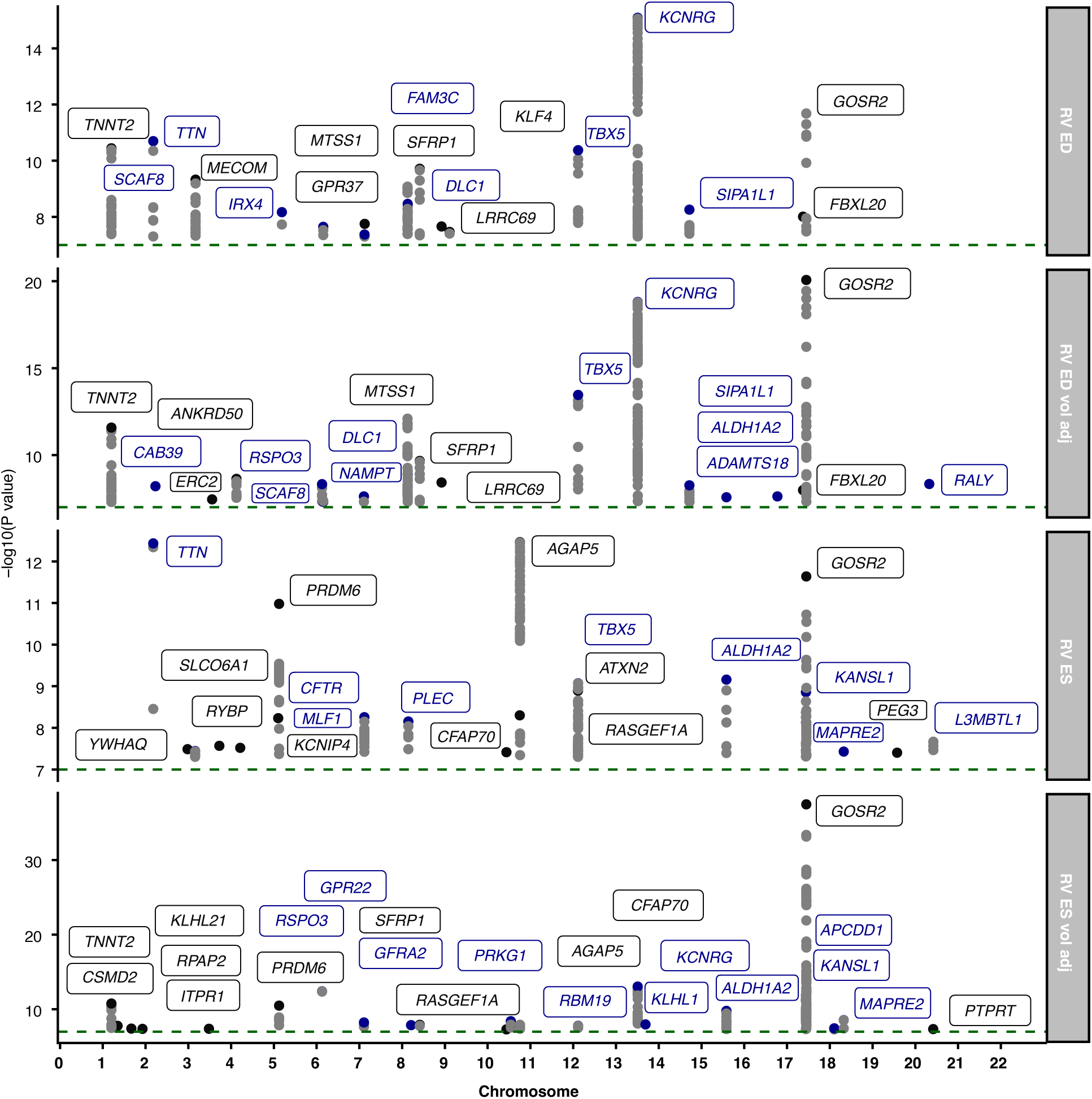
Manhattan plots of the statistically significant genetic loci associated with right ventricular trabeculation. Common variants were identified from genome-wide association studies of trabecular complexity assessed with fractal dimension (FD) analysis. The prioritised gene is noted for the statistically significant loci identified through common variant GWAS of four meta-Manhattan plots of the minimum P value for each SNP across measures of trabeculation: RV ED, right ventricular measures of FD in end-diastole; RV ED vol adj, right ventricular measures of FD in end-diastole and adjusted for right ventricular end-diastolic volume; RV ES, right ventricular measures of FD in end-systole; RV ES vola adj, right ventricular measures of FD in end-systole and adjusted for right ventricular end-systolic volume. The GWAS results were statistically significant if P<5 × 10^−8^. Blue, a visual aid that changes with chromosomal position on the x-axis.

The genomic loci of *AGAP5, ATXN2*, and *CFTR* are unique associations to FD measures in the right heart. Variants in *ATXN2* cause spinocerebellar ataxia, a neurodegenerative condition with cardiac autonomic dysfunction. Variants in *CFTR* cause cystic fibrosis, a rare inherited genetic condition that causes breathing and digestive problems. AGAP5 has a GTPase-activating role, and the locus has been associated with cardiac features and heart failure previously^32,33^.

Associations in the loci of *PRDM6, GOSR2, ALDH1A2, AGAP5, KANSL1, TNNT2, MTSS1, SFRP1, TBX5, KCNRG*, and *SIPA1L1*, remained after adjustment for volume, which also identified additional loci (*CSMD2, KLHL21, KLHL1, GPR22, KCNRG, PRPRT, RASGEF1A, ANKRD50, DLC1*).

The SNP-based heritability was estimated for each of the trabecular morphology measures (**Figure S7**). Mean RV FD had the highest estimated heritability (h^2^=0.26-0.30). The slices in the mid-region of the RV (slices 4-5) had the highest estimated heritability (h^2^∼0.25).

### Rare variant association studies identified the burden of protein-altering variants

We identified 45 genes in which protein-altering variants were associated with RV trabecular morphology in 33,030 European participants with exome sequencing data (**Figure 7, Figure S8, Tables S3-S4**). This included genes with clinical relevance (clinicalgenome.org), implicated in rare syndromes, and those known to regulate trabeculation. Significant gene ontology (GO) resource enrichment analysis of the genes identified from the GWAS and this burden analysis showed the strongest relationship with A band, sarcomere, myofibril, and contractile muscle fibres (**Table S5**).

**Figure 7.**
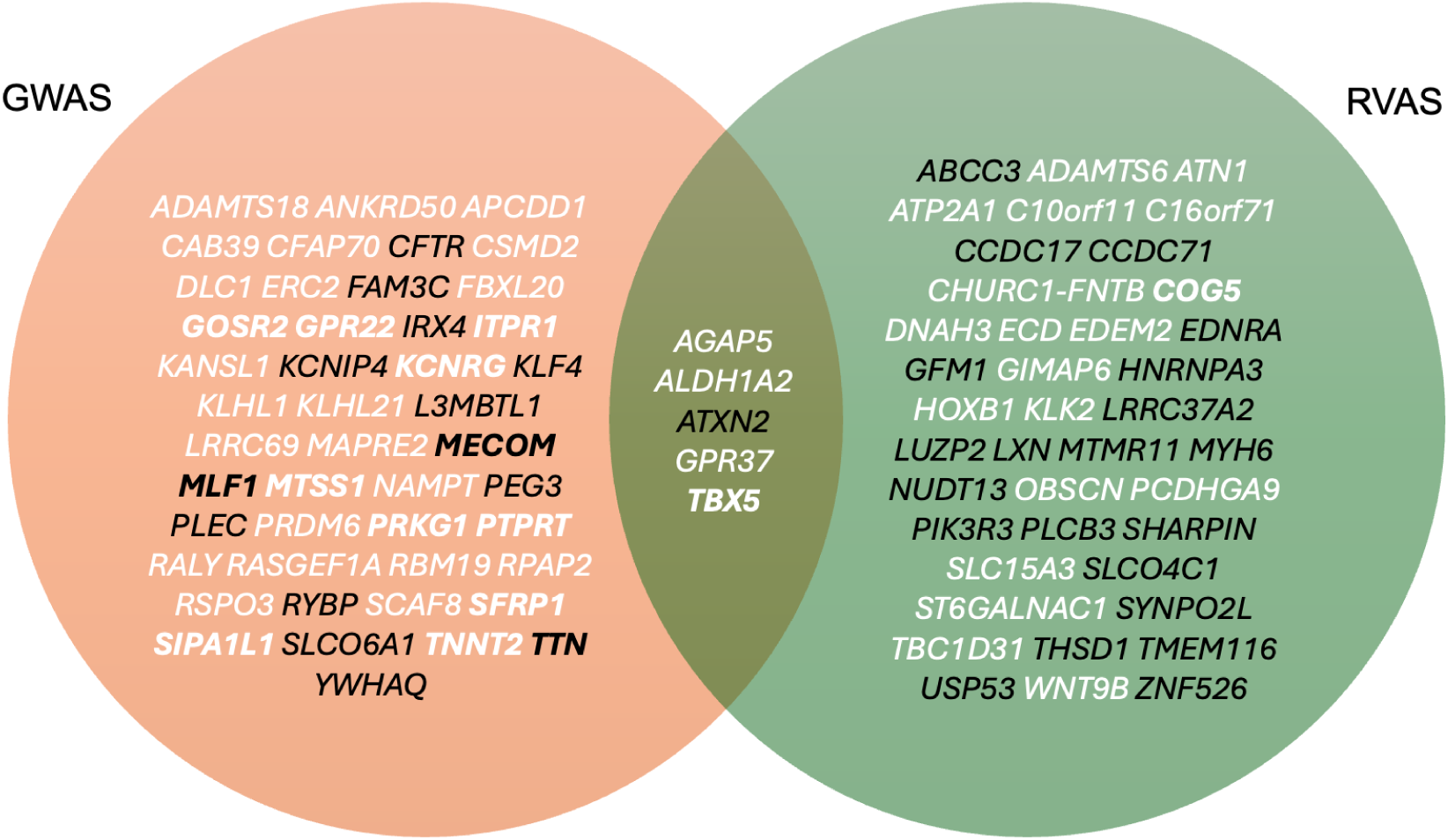
Venn diagram of the prioritised genes from the genetic association studies. Five genes were identified through common variant and protein-altering variant genetic association studies. Bold, 15 genes were found for LV trabeculation^17^, suggesting a bi-ventricular trabeculation modifier. White, genes associated with trabeculation when adjusted for ventricular volume. GWAS, genome-wide association studies; RVAS, rare variant association studies.

RV trabeculation was influenced by genes with roles in cardiac function and disease (*TBX5, MYH6*). Both *TBX5* and *MYH6* are associated with congenital heart disease, and septal defect associated with RV trabecular morphology at PheWAS (see **Figure 4**). T-box 5 (*TBX5*) is a transcription factor responsible for the septation of the heart and upper limbs during embryonic development, and variants in the gene can cause Holt-Oram syndrome (characterised by heart septal defects and conduction disorders). The locus was identified in a genetic analysis of LV trabecular morphology^17^, has been described for what is termed left ventricular noncompaction (LVNC) cardiomyopathy^3,18,30,31^, and was found here to associate with maximum RV FD in end systole, suggesting that it is a genetic modifier of the development of biventricular trabeculation, as well as trabecular structure and function.

Variants in myosin heavy chain 6 (*MYH6*) have definitive evidence for causing multiple types of congenital heart disease. The *MYH6* gene encodes the alpha heavy chain subunit of a fast ATPase primarily expressed in human atrial tissue (GTeXportal.org). The association here with adult trabecular morphology through volume in the basal portion of the RV (not previously identified for the LV) suggests that it is a modifier of cardiac structure beyond its involvement in overt congenital disease.

*SYNPO2L* (Synaptopodin 2-like) and *AGAP5* (ArfGAP With GTPase Domain, Ankyrin Repeat And PH Domain 5) associated with increased end-systolic measures of RV trabecular morphology. *SYNPO2L* is a protein-coding gene that acts as a Z-disc component in cardiac and skeletal muscles, crucial for actin filament bundling and sarcomere organisation and plays a key role in heart structure development. Variants in *SYNPO2L* and *AGAP5* are known genomic loci of atrial fibrillation and heart failure^33^. The gene of the obscurin protein (*OBSCN*) was identified; there is some evidence for the locus in cardiomyopathies, and it has been previously curated for LVNC^31^. COG5 (Component Of Oligomeric Golgi Complex 5) has a role in glycosylation disorders; the gene was also found to be associated with LV trabecular morphology^17^.

Three additional loci were found at both RV FD GWAS and burden analyses (*ALDH1A2, GPR37* and *ATXN2*). *ATXN2* was also found in GWAS. ALDH1A2 (Aldehyde Dehydrogenase 1 Family Member A2) catalyses the synthesis of retinoic acid (vitamin A) from retinaldehyde, with an important role in cardiac development. *GPR37* (G protein-coupled receptor 37) is mainly expressed in the brain and has been linked to Parkinson’s disease and lung adenocarcinoma.

Overall, these findings emphasise that RV trabeculation is influenced by a diverse set of genes involved in cardiac development, structural integrity, and disease susceptibility, and by enrichment in pathways related to sarcomere structure and contractile muscle components.

## Discussion

Population-level studies of myocardial trabeculae have, to date, been predominantly LV-centric. Yet RV structure and function are strong predictors of pulmonary disease, congenital heart disease, arrhythmias, and heart failure^34^. Although the RV and LV are closely interrelated through systolic and diastolic interdependence^34^, the extent to which RV trabeculation may serve as a biomarker of cardiac remodelling, disease, and adaptation, and whether it can identify genetic modifiers of cardiac structure and function beyond those of the LV, is unknown.

Here, we analysed RV myocardial trabecular traits in adult humans across the allele frequency spectrum, identifying new loci that regulate complex biological patterning and conduction signalling. We characterised the genetic modifiers of RV trabeculation, including loci with bi-ventricular roles and others with roles specific to the RV in congenital heart disease and respiratory conditions. The replicated genomic loci of *TBX5, TNNT2, GOSR2, KCNRG, TTN, SFRP1, MECOM, MTSS1, ITPR1, MLF1, COG5, GPR22, PRKG1, PTPRT* and *SIPA1L1* have now been identified across sample sizes and ventricles as influencing whole-heart trabecular morphology^6,17^. Together, they constitute a network implicated in cardiac development, electrophysiology, myocardial structure, and cardiac remodelling, with roles spanning transcriptional regulation, calcium handling, cell adhesion, cytoskeletal organisation, and intracellular signalling.

38 common variant loci and 44 genes from burden testing were independent of previous studies of trabecular morphology of the LV in end-diastole. Newly identified genomic loci at *CFTR* and *MYH6*, together with phenome-wide association analyses of RV trabecular morphology, reveal that the RV has distinct relationships with respiratory conditions, consistent with its coupling to the pulmonary circulation, as well as with congenital heart disease. The identification of *CFTR* opens a novel mechanistic avenue for understanding cardiac stressors, disease development, and ventricular remodelling through modifiers of lung and metabolic function. As a chloride and bicarbonate ion channel highly expressed in airway, pancreatic, and intestinal epithelia, its association with RV trabeculation may reflect an adaptive response to right heart strain and respiratory compliance. The identification of *MYH6* complements our understanding of adult outflow tract-linked ventricular patterning, implicating either developmental timing and sarcomeric control of cardiac morphology, or the influence of atrial genes on right ventricular structure and function.

Among the range of conditions assessed, RV imaging-derived measures showed the strongest associations with diabetes, both with and without adjustment for ventricular volume. A prior study of RV imaging-derived traits in 340 diabetic patients identified smaller RV volumes, hyperdynamic systolic function, and impaired relaxation relative to controls^35^. These findings suggest that systemic metabolic dysfunction, linked to ventricular stiffening and microvascular disease, affects not only loading conditions and remodelling but also myocardial trabecular adaptation, consistent with evidence of reduced contractility in trabeculae from diabetic rat hearts^36^. The association between diabetes and altered RV traits also has implications for physical activity, as RVEDV and RVSV were independent determinants of peak VO_2_ on cardiopulmonary exercise testing in the same diabetic cohort^35^.

RV hypertrabeculation has been described in “isolation” with right heart failure, with one report implicating a *TTN* missense variant^37^. The variant, described as “NM_001256850: c.16799T>C: p.L5525S” is not identifiable by current annotation and most likely refers to c.16574T>C (MANE Select: NM_001267550.2: c.17525T>C: p.L5842S) located in a *TTN* exon with low cardiac expression^38^. Only a small number of *TTN* missense variants have been reported as pathogenic, and robust evidence, including segregation data, is required to implicate them in cardiomyopathy, given their prevalence across the gene in the general population. We have previously discussed the longstanding diagnostic ambiguity surrounding LV non-compaction^18^, and we similarly caution against using RV trabecular morphology alone as a diagnostic criterion. Rather, it should be considered a useful biomarker of cardiac remodelling that is shaped by genetic factors associated with disease.

A limitation of the analysis is single-time-point data, so we do not know the temporal trajectory of trabecular adaptation and the relationship with remodelling. Repeated imaging data is being released for UK Biobank participants. UK Biobank is a large cross-sectional study that is subject to survival bias and latent population stratification; however, risk factor associations appear to be broadly generalisable and are statistically powered^39^.

In conclusion, our findings demonstrate that genetic variation spanning rare and common alleles influences right ventricular trabecular architecture in the adult human heart, engaging biological pathways involved in early cardiac morphogenesis, lineage specification, and cytoskeletal organisation. The identified loci further connect RV trabeculation to congenital heart disease and pulmonary pathology, alongside factors previously identified for the left ventricle. This complex and adaptable structural feature of the myocardium emerges as a phenotypic readout of broad developmental and disease-relevant processes, with relevance to cardiovascular health across the life course.

## Data availability

All data arising from this analysis will be made available through the UK Biobank (https://biobank.ndph.ox.ac.uk/showcase/) and GWAS catalog (https://www.ebi.ac.uk/gwas/). UK Biobank data is publicly available on application (https://www.ukbiobank.ac.uk/).

## CrediT statement

Conceptualization: K.A.M., D.P.O’R.; Methodology: K.A.M.; Formal Analysis: K.A.M.; Resources: D.P.O’R., F.N., J.S.W.; Data Curation: K.A.M., J.S., R.J., A.S.; Writing – Original Draft: K.A.M.; Writing – Review & Editing: (all authors); Visualization: K.A.M., D.P.O’R; Funding Acquisition: K.A.M.

## Methods

### Study overview

The UK Biobank (UKB) study recruited 500,000 participants aged 40 to 69 years old from across the United Kingdom between 2006 and 2010 (National Research Ethics Service, 11/NW/0382)^20^. Genotyping array data and exome sequencing data are available for over 450,000 participants. This study was conducted under the terms of access to projects 40616 and 47602. All participants provided written informed consent. A sub-study recalled participants for cardiac magnetic resonance imaging (CMR)^21^, and volumetric traits were measured using quality-controlled deep learning algorithms^22^.

### Fractal analysis of trabecular morphology

We optimised our automated fractal analysis so that it is suitable for biobank-scale analyses of right ventricle geometry at both end-diastole and end-systole^26^. Convolutional neural networks were used for short-axis cine end-diastolic and end-systolic right ventricular segmentation to label pixels containing myocardium. The performance of image annotation using this algorithm is equivalent to a consensus of expert human readers and achieves subpixel accuracy for cardiac segmentation^22,26^. Each image slice was cropped to the maximum extent of the analysed RV mask plus 10mm in all directions to include the adjacent myocardium, preserving the original labels. The cropped images were interpolated to 0.25×0.25mm pixels with bicubic interpolation. The pixel intensities within the heart were normalised. The level-set method^40^ was used to segment the blood pool and trabeculae within the ventricle label to identify the trabecular boundary. The outline of the resulting blood pool segment was obtained by Sobel edge-detection. The fractal dimension was calculated from the smallest box-counting size of 2 pixels and the largest size of 25% of the shorter image side to assess the negative slope of the double logarithmic curve of the box size versus the box count. For the standard box-counting method, the image is overlain by repeated grids of known box size, and the number of boxes containing non-zero image pixels is recorded. Fractal analysis was automated using FracAnalyse software^5,41^ and adapted for the scale of the UK Biobank and derived segmentations (AutoFD^6,17,26^). The analysis was performed on each right ventricular slice. We did not differentiate between trabeculae and papillary muscles, as edge detection was applied within the whole LV cavity.

Fractal dimension (FD) is defined as the negative gradient of an ordinary least-squares fit line to the logarithm of box size and box count. FD is a scale-invariant ratio providing an index of complexity. The raw FD measures were interpolated using a Gaussian kernel local fit to a nine-slice template to allow for comparison across subjects in end-diastole, and seven slices for end-systole. Summary statistics (minimum, mean, median, maximum) were calculated from the slices. The measures were adjusted for covariates: age at scan, age^2^, sex, age:sex interaction, imaging centre, imaging release, body surface area, systolic blood pressure, days per week of vigorous exercise, and 10 genetic principal components for ancestry, through multiple linear regression and the resulting standardized residuals (mean=0, SD=1) were normalized by an inverse rank normalization.

### Electrocardiogram-derived diagnoses

The electrocardiograms (ECGs) were performed according to a defined protocol and analysed using proprietary software (GE CardioSoft, Boston, MA). Data from the first imaging visit (instance 2, n=42,386) was labelled using a previously trained convolutional neural network designed to identify six diagnoses from the ECG^29^: sinus bradycardia, sinus tachycardia, left bundle branch block, right bundle branch block, 1st-degree AV block, and atrial fibrillation. The binary outputs (presence or absence of each diagnosis) were used for subsequent analyses. Automated diagnoses had F1 scores above 80% and specificity over 99% and were previously shown to be superior to cardiology resident medical doctors^29^. The ECGs were preprocessed with a bandpass filter of 0.5 to 100hz, a notch filter at 60hz, and resampling to 400hz. Zero-padding resulted in a signal with 4,096 samples per lead for a 10s recording, which was used as input to a neural network model.

### Genetic association studies

Genotype calling was performed on the UK BiLEVE Axiom array and the UK Biobank Axiom array, resulting in 805,426 markers in GRCh37 coordinates and quality control was undertaken^20^. The dataset was phased, and 96M genotypes were imputed using the Haplotype Reference Consortium and UK10K haplotype resources^20^. Exome sequencing was undertaken on the Illumina NovaSeq 6000 platform. Reads were mapped to the hg38 reference genome, and quality control was undertaken^42^.

For genome-wide association studies (GWAS), the imputed UK Biobank genotyping data were converted from bgen format to plink. Minor allele frequency of >0.001 in autosomes was included. Individuals with more than 5% missing genotypes and SNPs with more than 5% missingness were excluded. Participant sex discrepancies, heterozygosity, and relatedness were handled by keeping only European individuals and participants included in the UKBB principal components analysis^6,20^. SNPs deviating from Hardy-Weinberg equilibrium (1×10^−8^) and those with an imputation INFO score of <0.4 were excluded. Individuals with trabeculation data were extracted, and GWAS was undertaken for 34,110 participants using GCTA software (version 64). A sparse genetic relationship matrix (GRM) was created, and FastGWA was undertaken with a mixed linear model, adjusting for the genotyping array batch. The genes of the significant GWAS results (*P*P<5 × 10^−8^) were prioritised through LocusZoom and eQTLs from GTEx v8. Phenotype associations through GWAS catalog, ClinGen, and PheWEB, were assessed, and resulting genes were analysed by gene ontology (GO) resource enrichment analysis using Panther.

Rare variant association studies (RVAS) (protein-altering burden analyses) were undertaken using Regenie software on the DNA Nexus Research Analysis Platform. The raw genotyping data for step 1 of Regenie were lifted from GrCh37 to GrCh38. SNPs in autosomes with a minor allele frequency <0.01, missingness of >0.01, a minor allele count of <20, deviations from Hardy-Weinberg equilibrium (5×10^−15^), and individuals with greater than 10% missingness, were excluded from the analysis. Interchromosome, SNPs in linkage disequilibrium (indep-pairwise 1000 100 0.9), and areas of low complexity, were excluded for step 1. Exome sequencing data for step 2 were quality controlled for variants in the autosomes with missingness less than 10%, variants where less than 90% of all genotypes for that variant had a read depth less than 10, deviations from Hardy-Weinberg equilibrium (1×10^−15^), and individuals with more than 10% missingness. Step 2 of Regenie was run for the trabeculation traits over different allele frequencies (singletons, 0.01, 0.001) for 6 overlapping, protein-altering variant, custom masks (LoF only; missense only (flagged by >1 of 5 deleterious software); missense only (all); missense only (flagged by all of 5 deleterious software); protein-altering variants (LoF and missense flagged by >1 of 5 deleterious software); protein-altering variants (all Lof and missense)), where the minimum minor allele count was at least 3. Bonferroni significance for 18,117 included genes was *P*<2.76×10^−6^.

### Statistical analysis

The Student’s t-test was used to assess differences in means. Pearson’s correlation coefficient described the relationships. Effect sizes are presented as standardised beta coefficients. Phenome-wide association studies (PheWAS) were undertaken using the PheWAS R package with clinical outcomes and coded phenotypes converted to 1,840 categorical PheCodes. P-values were deemed significant with Bonferroni adjustment for the number of PheCodes. Heritability was estimated by creating a genetic relationship matrix in GCTA and using a restricted maximum likelihood analysis (REML) to estimate the variance explained by the SNPs that were used to estimate the GRM.

## Supplementary figures

Supplementary tables S1-S5 are presented in the file TrabecRVsuppl.xlsx.

**Supplementary Figure 1.**
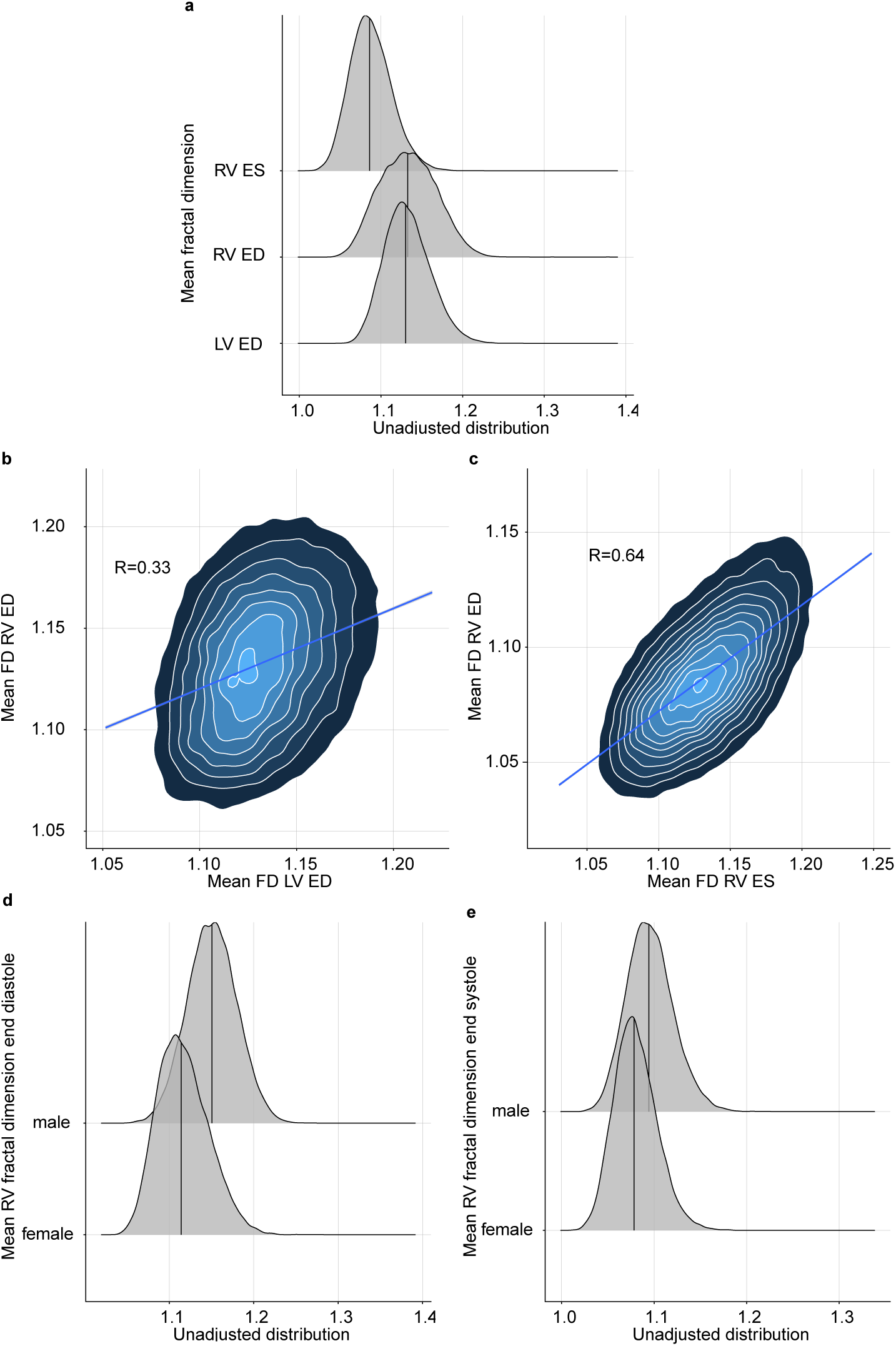
Summary comparisons of the fractal dimension analysis. **a**, The distribution of mean measures of fractal dimension by ventricle (RV, right; LV, left) and cardiac cycle phase (ED, end-diastole; ES, end-systole). b, The relationship between right (y-axis) and left (x-axis) mean trabecular morphology in end-diastole. c, The relationship between right ventricular mean trabecular morphology in end-diastole (y-axis) and end-systole (x-axis). d-e, The distribution of measures of fractal dimension by sex (d, end-diastole; e, end-systole). FD, fractal dimension; RV, right ventricle; ED, end-diastole; LV, left ventricle; ES, end-systole; R, Correlation coefficient.

**Supplementary Figure 2.**
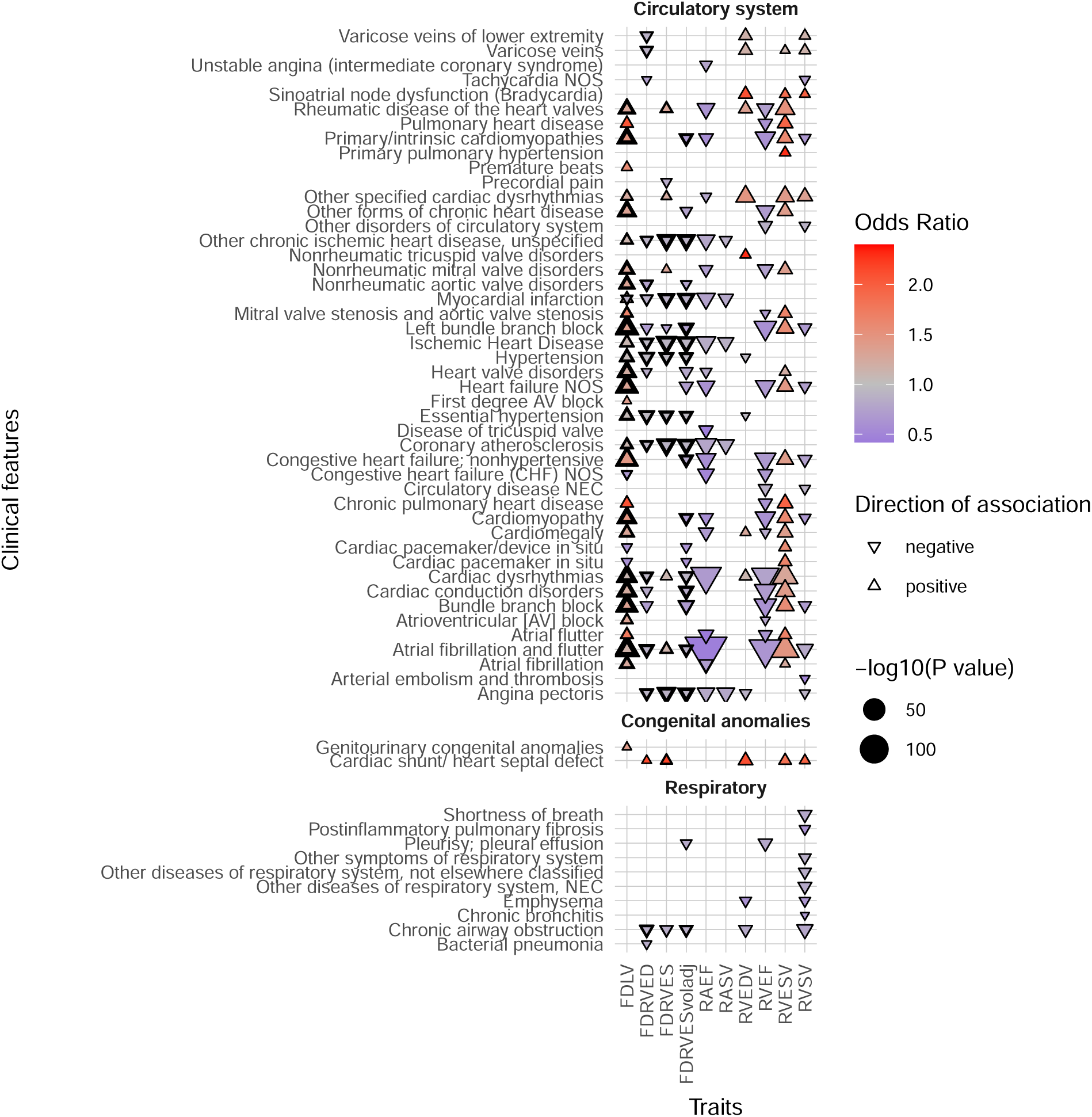
Phenome-wide association study of right ventricular trabecular complexity and other CMR-derived traits for the circulatory system, congenital anomalies, and respiratory categories of phenotypes. Trabecular morphology was analysed through fractal dimension analysis (FD) of the right ventricle (RV) in end-diastole (ED) and end-systole (ES). ED and ES volume-adjusted RV FD traits are presented separately (vol adj). FD of the left ventricle (LV FD) was added for comparison. Right ventricular end-diastolic volume (RVEDV), end-systolic volume (RVESV), ejection fraction (RVEF), stroke volume (RVSV), atrial ejection fraction and stroke volume (RAEF, RASV), were analysed to assess for pleiotropy between trabecular complexity (FD) and remodelling traits altered in cardiac disease. Fractal dimension was measured at different spatial locations in the right ventricle, and the aggregate of all results is shown. The analyses were completed on 45,681 participants of the UK Biobank population. Phenotypes as phecodes are described on the y-axis, with the phecode category separating the groups and the imaging traits are on the x-axis. Each point denotes a significant PheWAS association with a Bonferroni correction for 1,163 analysed phecodes. The shape and colour denote the direction of effect and the odds ratio.

**Supplementary Figure 3.**
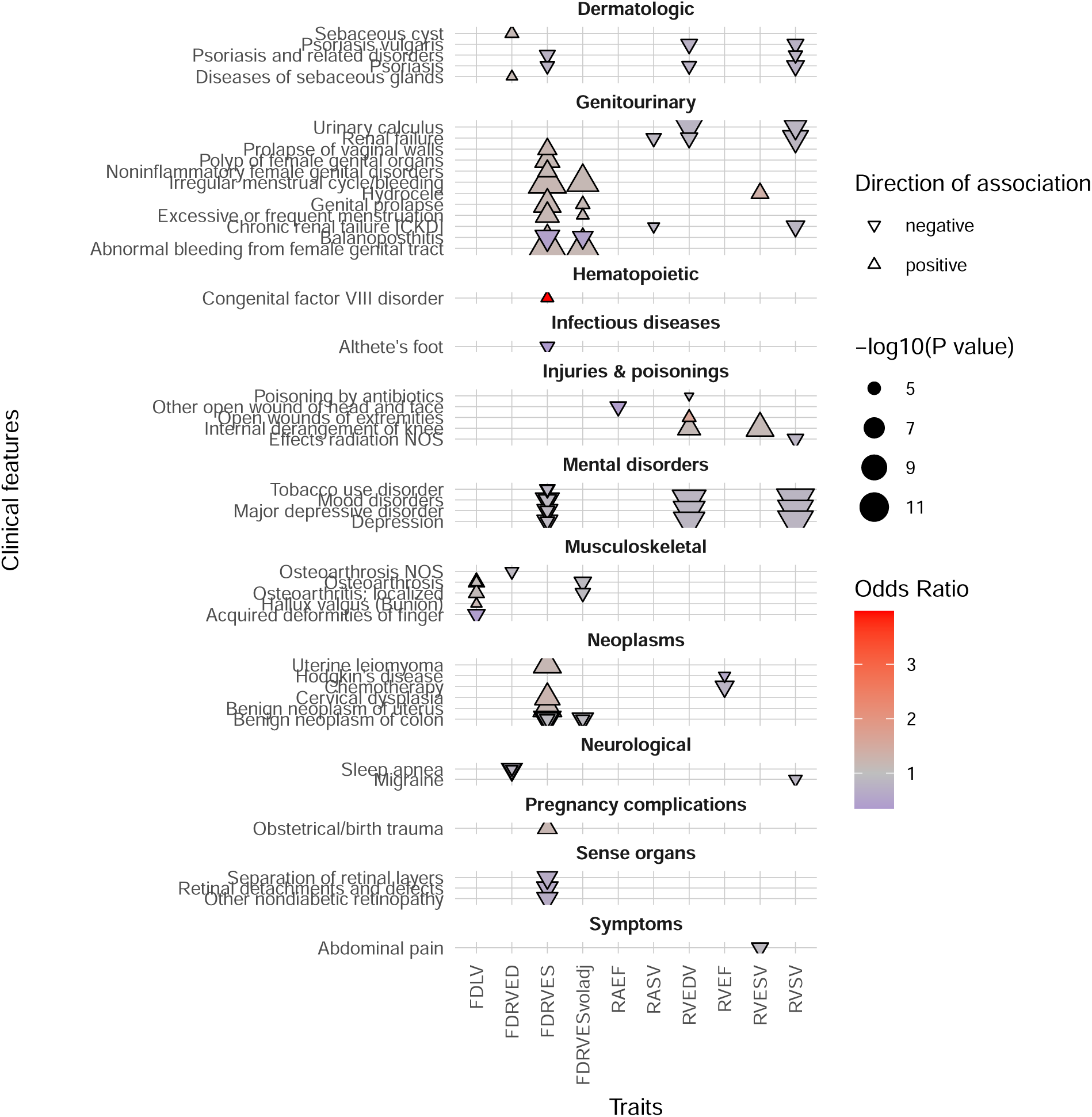
Phenome-wide association study of right ventricular trabecular complexity and other CMR-derived traits for a range of categories of phenotypes. Trabecular morphology was analysed through fractal dimension analysis (FD) of the right ventricle (RV) in end-diastole (ED) and end-systole (ES). ED and ES volume-adjusted RV FD traits are presented separately (vol adj). FD of the left ventricle (LV FD) was added for comparison. Right ventricular end-diastolic volume (RVEDV), end-systolic volume (RVESV), ejection fraction (RVEF), stroke volume (RVSV), atrial ejection fraction and stroke volume (RAEF, RASV), were analysed to assess for pleiotropy between trabecular complexity (FD) and remodelling traits altered in cardiac disease. Fractal dimension was measured at different spatial locations in the right ventricle, and the aggregate of all results is shown. The analyses were completed on 45,681 participants of the UK Biobank population. Phenotypes as phecodes are described on the y-axis, with the phecode category separating the groups and the imaging traits are on the x-axis. Each point denotes a significant PheWAS association with a Bonferroni correction for 1,163 analysed phecodes. The shape and colour denote the direction of effect and the odds ratio.

**Supplementary Figure 4.**
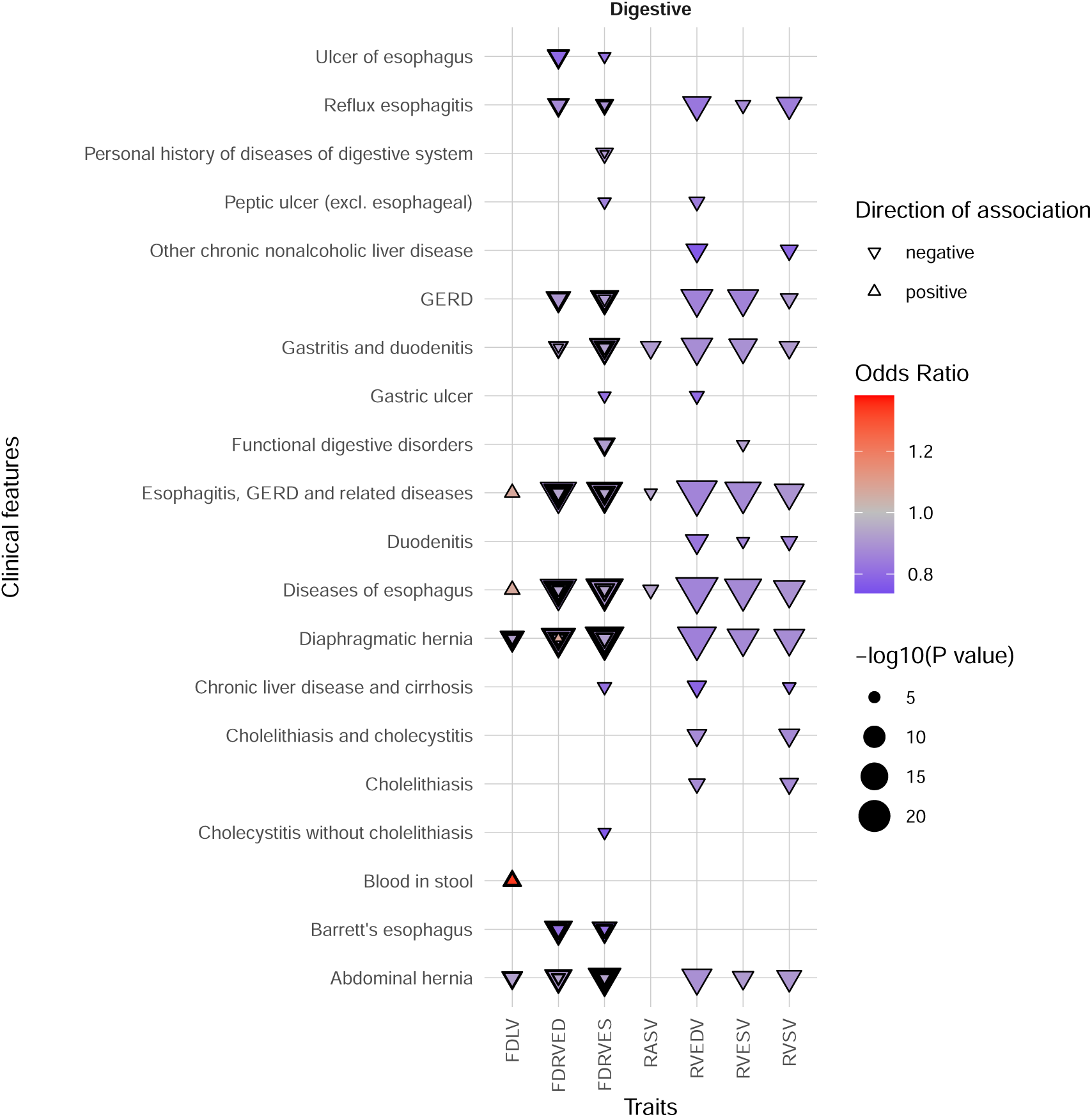
Phenome-wide association study of right ventricular trabecular complexity and other CMR-derived traits for the digestive category of phenotypes. Trabecular morphology was analysed through fractal dimension analysis (FD) of the right ventricle (RV) in end-diastole (ED) and end-systole (ES). ED and ES volume-adjusted RV FD traits are presented separately (vol adj). FD of the left ventricle (LV FD) was added for comparison. Right ventricular end-diastolic volume (RVEDV), end-systolic volume (RVESV), ejection fraction (RVEF), stroke volume (RVSV), atrial ejection fraction and stroke volume (RAEF, RASV), were analysed to assess for pleiotropy between trabecular complexity (FD) and remodelling traits altered in cardiac disease. Fractal dimension was measured at different spatial locations in the right ventricle, and the aggregate of all results is shown. The analyses were completed on 45,681 participants of the UK Biobank population. Phenotypes as phecodes are described on the y-axis, with the phecode category separating the groups and the imaging traits are on the x-axis. Each point denotes a significant PheWAS association with a Bonferroni correction for 1,163 analysed phecodes. The shape and colour denote the direction of effect and the odds ratio.

**Supplementary Figure 5.**
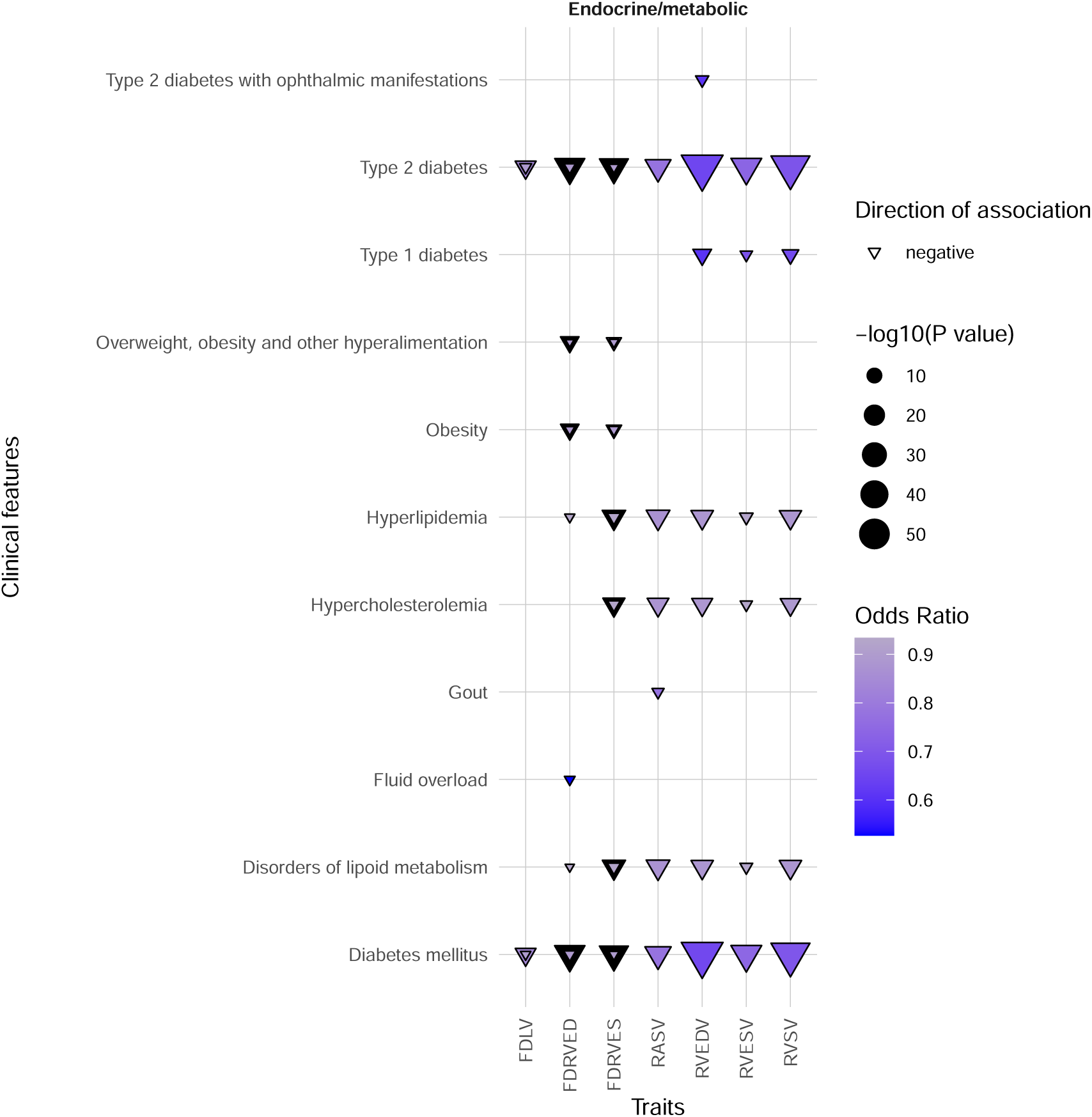
Phenome-wide association study of right ventricular trabecular complexity and other CMR-derived traits for the endocrine/metabolic category of phenotypes. Trabecular morphology was analysed through fractal dimension analysis (FD) of the right ventricle (RV) in end-diastole (ED) and end-systole (ES). ED and ES volume-adjusted RV FD traits are presented separately (vol adj). FD of the left ventricle (LV FD) was added for comparison. Right ventricular end-diastolic volume (RVEDV), end-systolic volume (RVESV), ejection fraction (RVEF), stroke volume (RVSV), atrial ejection fraction and stroke volume (RAEF, RASV), were analysed to assess for pleiotropy between trabecular complexity (FD) and remodelling traits altered in cardiac disease. Fractal dimension was measured at different spatial locations in the right ventricle, and the aggregate of all results is shown. The analyses were completed on 45,681 participants of the UK Biobank population. Phenotypes as phecodes are described on the y-axis, with the phecode category separating the groups and the imaging traits are on the x-axis. Each point denotes a significant PheWAS association with a Bonferroni correction for 1,163 analysed phecodes. The shape and colour denote the direction of effect and the odds ratio.

**Supplementary Figure 6.**
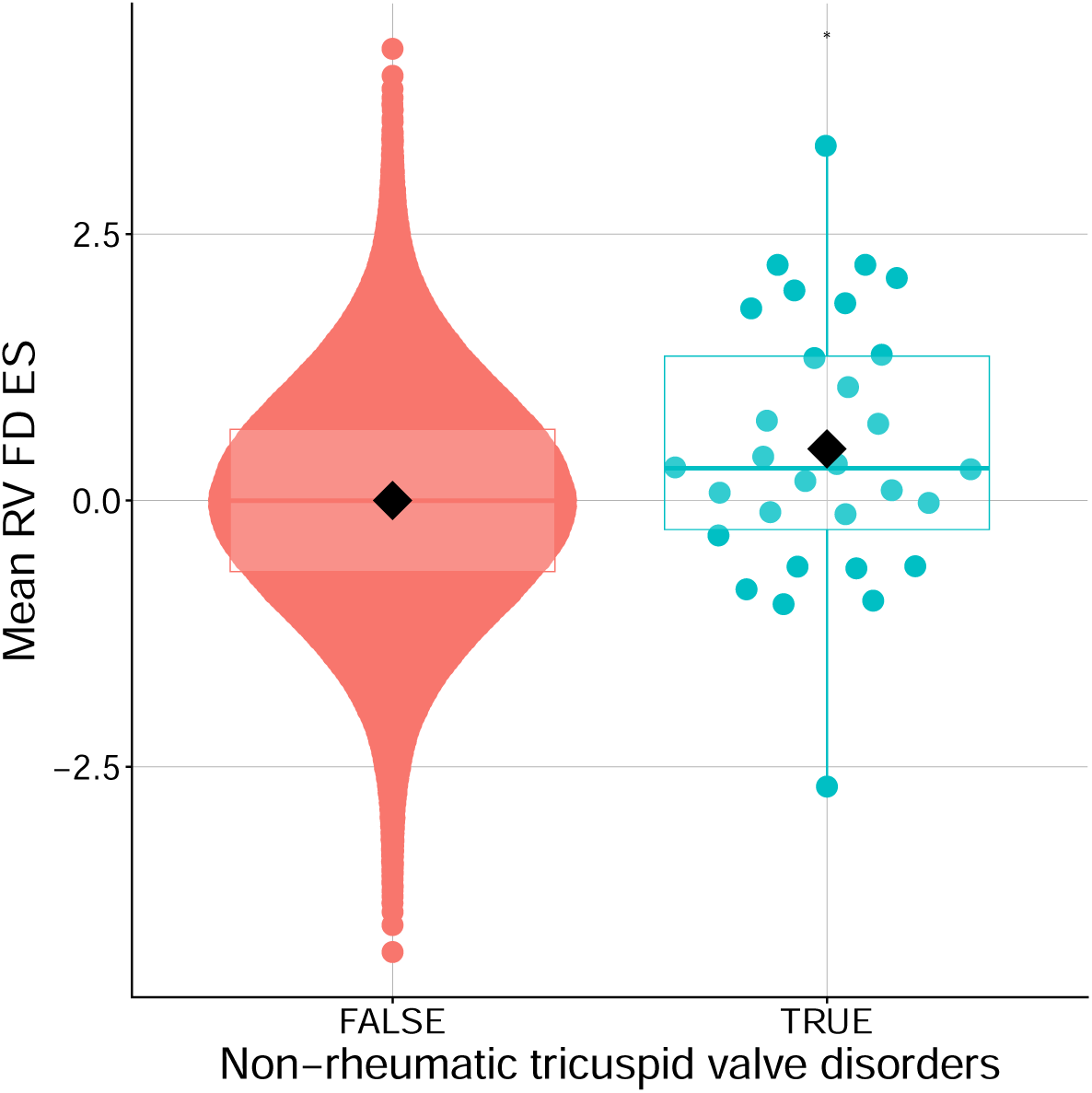
Individuals diagnosed with tricuspid valve disease had increased mean right ventricular fractal dimension in end-systole. The plot depicts diagnosed individuals (x-axis=TRUE) with non-rheumatic tricuspid valve disease compared to non-diagnosed individuals (x-axis=FALSE). Mean right ventricular trabeculation measured in end-systole is presented on the y-axis. A nominal association was observed via Student’s t-test, but it is underpowered (ICD code 36, n=30 diagnosed in 42,850, P=0.04). FD, fractal dimension; RV, right ventricle; ES, end-systole; *, P=0.04.

**Supplementary Figure 7.**
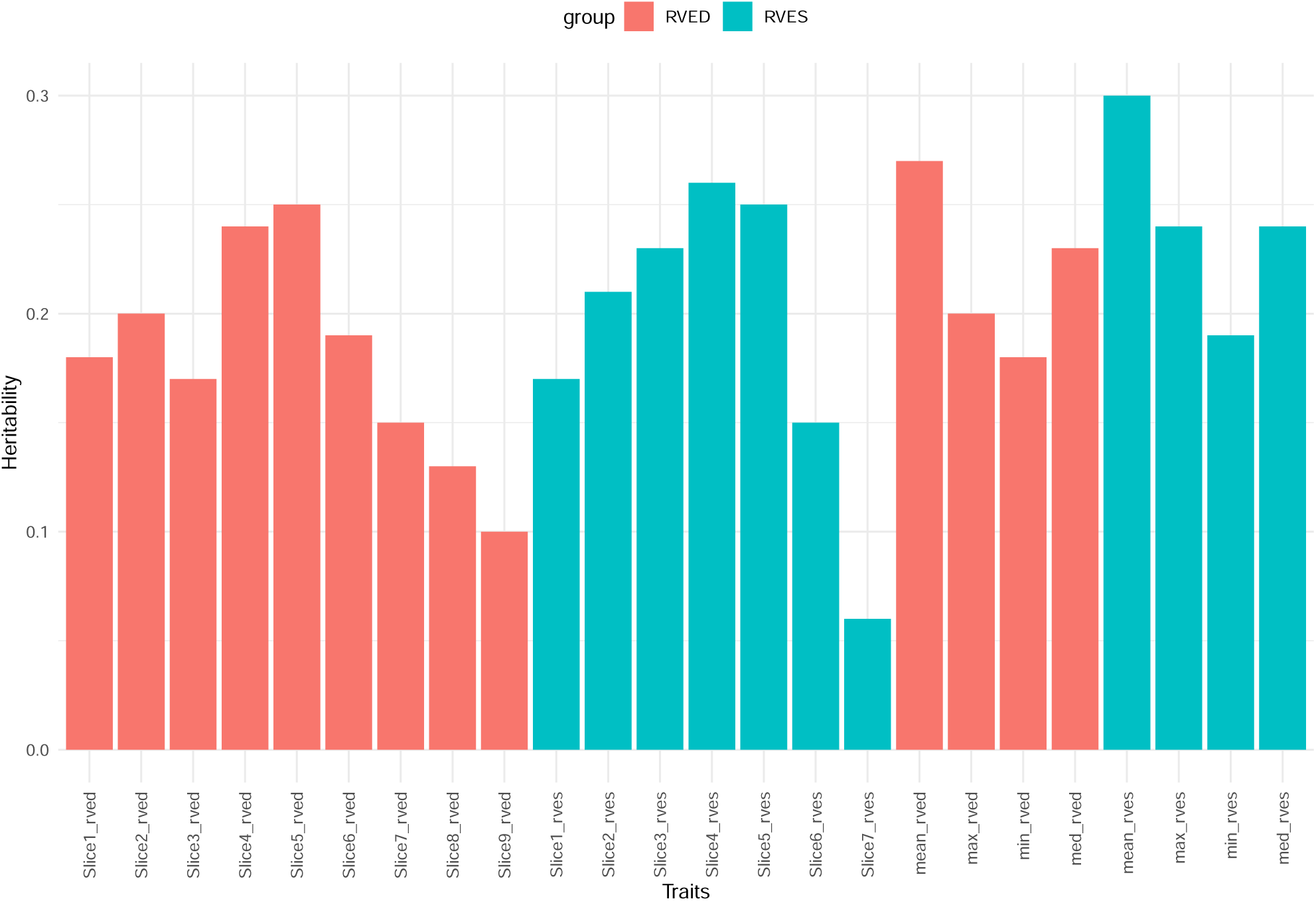
Estimated narrow-sense heritability (h^2^) for each fractal dimension trait. Heritability, the amount of influence that genetic factors have over a trait, was between 5%-30% for right ventricular (RV)-derived trabecular morphology. The plot depicts traits measured in end-diastole (ED) in orange and end-systole (ES) in blue. Nine cine slices were analysed for ED and seven for ES. Summary measures (min, minimum; max, maximum; mean; med, median) are also presented.

**Supplementary Figure 8.**
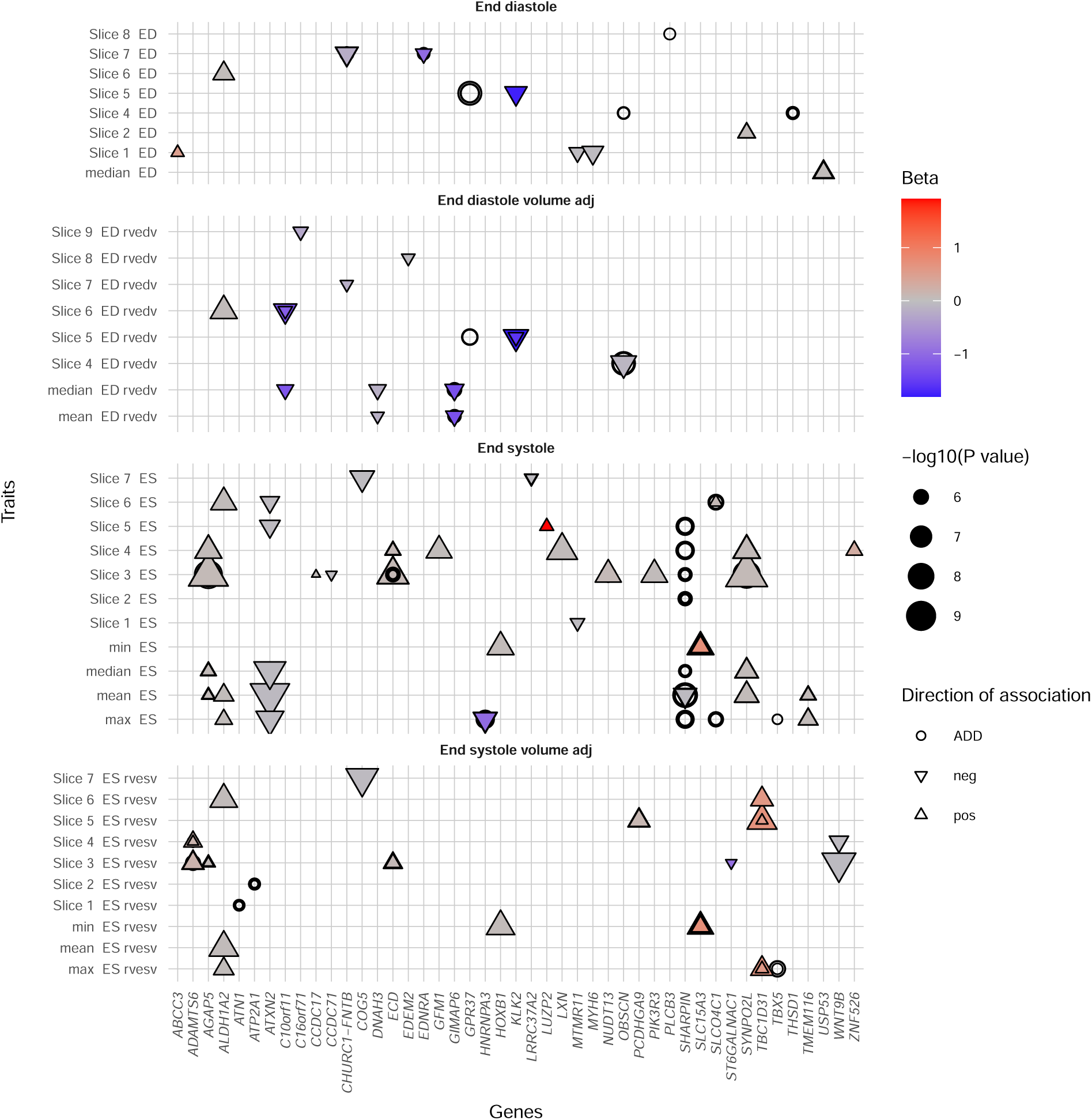
Plot for genes with a statistically significant burden of protein-altering variants associated with right ventricular trabeculation. Rare variant genetic analyses for trabecular complexity assessed with fractal dimension analysis. The gene is noted for the statistically significant group of protein-altering variants that associated with trabecular morphology. The results were statistically significant after adjusting for the number of genes analysed. Betas for the SKAT, SKATO, ACATV and ACATO (described as the “ADD” group), p-value only tests were denoted as 0 and presented as a circle for inclusion in the plot. ES, end-systole; ED, end-diastole; rvedv, adjusted for ED volume; rvesv, adjusted for ES volume; min, minimum; max, maximum; neg, negative; pos, positive; ADD, additional association testing providing p-values only.

